# Factors associated with differential seropositivity to *Leptospira interrogans* and *Leptospira kirschneri* in a high transmission urban setting for leptospirosis in Brazil

**DOI:** 10.1101/2023.04.10.23288388

**Authors:** Daiana de Oliveira, Hussein Khalil, Fabiana Almerinda G. Palma, Roberta Santana, Nivison Nery, Juan C. Quintero-Vélez, Caio Graco Zeppelini, Gielson Almeida do Sacramento, Jaqueline Cruz, Ricardo Lustosa, Igor Santana Ferreira, Ticiana Carvalho-Pereira, Peter J Diggle, Elsio A. Wunder, Albert I. Ko, Yeimi Alzate Lopez, Mike Begon, Mitermayer G. Reis, Federico Costa

## Abstract

**Background:** Leptospirosis is a zoonosis caused by pathogenic species of bacteria belonging to the genus *Leptospira*. Most studies infer the epidemiological patterns of a single serogroup or aggregate all serogroups to estimate overall seropositivity, thus not exploring the risks of exposure to distinct serogroups. The present study aims to delineate the demographic, socioeconomic and environmental factors associated with seropositivity of *Leptospira* serogroup Icterohaemorraghiae and serogroup Cynopteri in an urban high transmission setting for leptospirosis in Brazil.

**Methods/Principal Findings:** We performed a cross-sectional serological study in five urban informal communities in the city of Salvador, Brazil. During the years 2018, 2020 2021, we recruited 2.808 residents and collected blood samples for serological analysis using microagglutination assays. We used a mixed-effect multinomial logistic regression model to identify risk factors associated with seropositivity for each serogroup. Seropositivity to Cynopteri increased with age in years (OR 1.03; 95% CI 1.01-1.06) and was higher in those living in houses with unplaster walls (exposed brick) (OR 1.68; 95% CI 1.09-2.59) and where cats were present near the household (OR 2.00; 95% CI 1.03-3.88). Seropositivity to Icterohaemorrhagiae also increased with age in years (OR 1.02; 95% CI 1.01-1.03) but was higher in males (OR 1.51; 95% CI 1.09-2.10), in those with work-related exposures (OR 1.71; 95% CI 1.10-2.66) or who had contact with sewage (OR 1.42; 95% CI 1.00-2.03). Spatial analysis showed differences in distribution of seropositivity to serogroups Icterohaemorrhagiae and Cynopteri within the five districts where study communities were situated.

**Conclusions/Significance:** Our data suggests distinct epidemiological patterns associated with serogroups Icterohaemorrhagiae and Cynopteri within the high-risk urban environment for leptospirosis and with differences of spatial niches. Future studies must identify the different pathogenic serogroups circulating in low-income areas, and further evaluate the potential role of cats in the transmission of the serogroup Cynopteri in urban settings.

## Introduction

Leptospirosis is a potentially severe infectious disease that affects animals (both domestic and wild) and humans [1]. The disease is caused by pathogenic species of the genus *Leptospira* that causes a wide array of clinical manifestations in humans [2]. Symptoms can range from asymptomatic infection and light fever to severe manifestations with risk of death [3]. Over a million cases occur globally every year, causing over 60 thousand deaths [4]. The majority of the disease burden occurs in developing nations that lack adequate infrastructure and health services [5]. Human infection occurs through direct contact with urine or contaminated secretions from infected animals, or indirectly through contact with contaminated soil and water [1,6].

The genus *Leptospira* comprises 69 species and over 300 serovars, classified in more than 20 serogroups [7–9]. The microscopic agglutination test (MAT) is the golden standard serodiagnosis for leptospirosis and can indicate the presumptive infecting serogroup [10]. However, isolation or specific laboratory techniques like DNA sequence analysis are needed for definitive serotyping [11]. Some serovars can infect multiple animal species while others are more adapted to specific animal hosts [2]. This host preference might explain why some serovars/serogroups are more frequent in certain geographic areas. The diversity of reservoirs, serovars and serogroups, together with environmental and socio-behavioral determinants, pose a challenge to the characterization of the epidemiologic patterns of leptospirosis. These factors can influence the risk of transmission, and consequently, increase incidence and cause outbreaks [12].

Historically, tropical and subtropical nations have seasonal leptospirosis incidence peaks associated with heavy rainfall and flooding events during the rainy season [12–15]. The informal accelerated expansion of large urban centers, coupled with inadequate housing and sanitation, has resulted in environmental and social conditions highly favorable to disease hosts and pathogen transmission [16]. Over 33% of the world population live in urban poor communities [17], a figure around 28% in Brazil [18], where, because of the deep social inequalities, there is a direct correlation between vulnerable environments, disaster risks and public health problems [19,20]. Historically, in the city of Salvador, where most of the population is of black and mixed races, people with lower income to live in the peripheral areas of cities without access to basic rights, and where the prevalence of health problems such as leptospirosis is greater than in other areas of the city.

Studies performed in the past two decades in Salvador, Brazil, reported *Leptospira interrogans* serogroup Icterohaemorrhagiae serovar Copenhageni (strain Fiocruz L1-130) as the serogroup responsible for over 95% of the clinical cases recorded throughout the city [12], as well as asymptomatic infections in residents of the urban community of Pau da Lima within Salvador [21,22]. The animal reservoir of the serovar Copenhageni and serogroup Icterohaemorrhagiae is the brown rat (*Rattus norvegicus*), as shown by previous studies in the same community [23,24]. These studies identified the seroincidence of *Leptospira kirschneri* serogroup Cynopteri (strain 3522C) as the second highest in residents of urban communities [25]. However, the animal reservoir of this serogroups and the determinants of its transmission, remain unknown.

Thus, investigating the *Leptospira* serologic profile of the residents in urban communities, accounting for serogroup-specific risk factors, will contribute to a better understanding of the risk determinants of urban infection and inform the development of more specific control measures. The present study aims to identify demographic, socioeconomic, and environmental factors associated with seropositivity of *Leptospira interrogans* Icterohaemorrhagiae (strain Fiocruz L1-130) and *Leptospira kirschneri* Cynopteri (strain 3522C) in residents of five urban low-income communities in Salvador, Brazil.

## METHODS

### Ethics statement

The study was approved by the Ethics Committee of Fundação Oswaldo Cruz, Instituto Gonçalo Moniz (CAAE 45217415.4.0000.0040), the Ethics Committee for Research of Instituto de Saúde Coletiva (UFBA, 041 / 17-2.245.914.17-2.245.914) and the Institutional Review Board of Yale University (no. 2000031554).

### Study design

We performed a cross-sectional study in five urban poor communities (Pau da Lima (PL), Alto do Cabrito (AdC), Marechal Rondon (MR), Nova Constituinte (NC) and Rio Sena (RS) in the city of Salvador, Bahia, Brazil (Figure 1). In PL, we collected samples during the period of November 2020 to February 2021. Previous studies in the area indicated an annual leptospiral incidence of 37.8 infections per thousand [21]. In the four other communities, namely: AdC, MR, NC and RS, sampling occurred from April to September 2018. The communities are considered low-income, with precarious sanitation and infrastructure.

**Figure 1.**
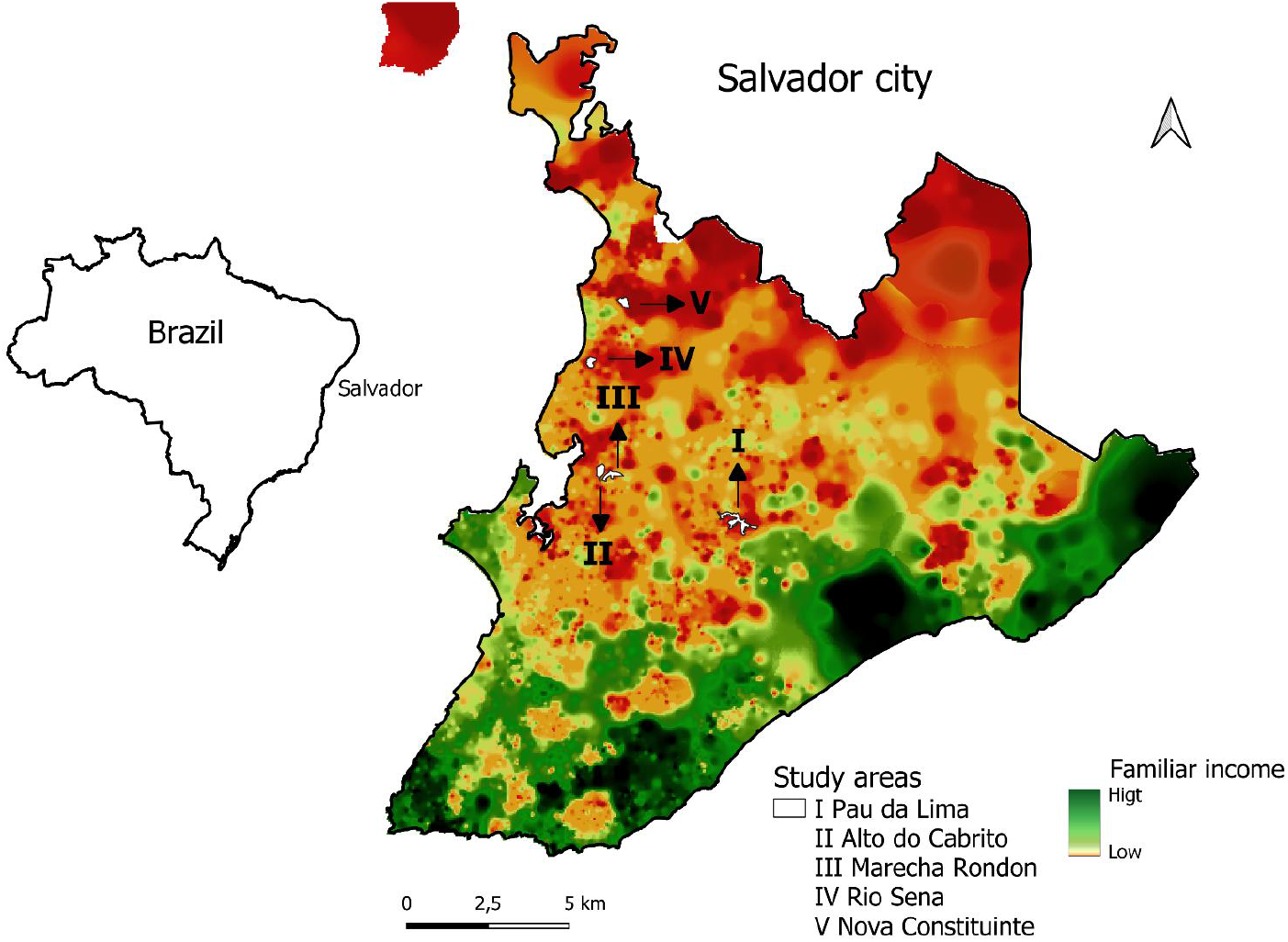
Urban communities in the city of Salvador, Brazil. Location of study sites (white) within the city. Distribution of family income, high (green) and low (orange).

Eligible individuals were recruited, and those willing to participate signed a free consent form and had a small blood sample drawn by a field professional. We applied a structured questionnaire to provide information on individual characteristics of the participant, domestic and peri-domestic environmental characteristics, exposure to sources of environmental contamination, and the presence of potential animal reservoirs.

Serological analysis was performed using the Microscopic Agglutination Test (MAT), the gold standard for human leptospirosis detection, as previously described [16]. MAT is based on dark-field microscopy detection of serum agglutination samples from an individual (antibodies) with live *Leptospira* antibodies. We performed association analysis only for serogroups Icterohaemorrhagiae strain Fiocruz L1-130 and Cynopteri strain 3522C, which were the most prevalent strains in the regions, according to our analyses.

### Outcomes, exposures, and covariates

The outcome of seropositive for leptospirosis was defined as seropositivity against serogroups Cynopteri and/or Icterohaemorrhagiae. A result was considered positive if 50% or more leptospires were agglutinated by MAT with a titer of ≥1:50. When agglutination was observed at a dilution of 1:100, the sample was titrated in serial two-fold dilutions to determine the highest positive titer [25].

The main explanatory variables were at the individual-level: age in years (as a quantitative variable), gender, ethnicity, highest education level achieved, receiving the Federal government stimulus program (Bolsa Família), employment status, work involving contact with sewer/garbage/construction materials, use of protective boots, contact with mud in the last 12 months, walking barefoot outside, and cleaning sewer canals in the last 12 months. In addition, environmental explanatory variables included proximity of the household to open sewers, paved access to the household, household with a backyard, plastered walls (proxy for quality of construction), garbage accumulation near the household, and presence of domestic animals (cats, dogs, and chickens).

### Spatial descriptive analysis

QGIS version 2.18.20 was used to construct a georeferenced database from satellite image WorldView-3 May 2017, at 31 cm resolution. The study team identified households within the study site and marked their positions onto hard copy 1:1,500 scale maps. The image WorldView-3 was acquired by the research project / Institute Gonçalo de Moniz - IGM - Fiocruz Bahia from the company Stamap, with disclosure permitted referencing the Copyrights of DigitalGlobe images. Kernel Density Estimation analysis (KDE) [26], with standard expression for two-dimensional spatial context: fhat(x) = n-1h-2 ∑k{(x-xi)/h}, where k(u) is the kernel function (a bivariate probability density), x1,…,xn are the data-points and h is the bandwidth, was performed to complementary analyses to evaluated possible territorial difference of niches between serogroups Icterohaemorrhagiae and Cynopteri with no overlapping patters territorial concentration. The study evaluates smoothed spatial distributions of sites with positive human cases of serogroups Icterohaemorrhagiae and Cynopteri, considering as a weight factor in the analysis the total number of cases in the household with distance matrix by 30 meters. The 30-meter distance matrix considered the average distance traveled by rats per day [27] as a possible exposure factor of households to serogroups, aiming to compare patterns of the Kernel analysis suitable for the scale of the maps in the evaluated areas. To determine the smoothed population-adjusted risk distribution we calculated the ratio of the KDE for positive cases of households to all households evaluated with residents tested by leptospirosis cases of serogroups Icterohaemorrhagiae and Cynopteri.

### Statistical Analysis

We performed a descriptive analysis using relative and absolute frequencies for qualitative variables and median and interquartile range for quantitative variables. A mixed-effect multinomial logistic regression model was used to estimate potential factors associated to seropositivity for both Icterohaemorrhagiae and Cynopteri serogroups. Variables included in the multivariate analysis were those with p < 0.15 in the bivariate analysis. The multivariable analyses were performed using the stepwise method based on the purposeful selection of variables, including both statistical and research criteria (biological plausibility) [21,28]. Also, -area” (five urban low-income communities) was include in the final model as a fixed effect. We checked for high levels of correlation among the predictor models that were selected to be included in the final model using Variance Inflation Factor (VIF), but VIF values were <5 and thus no variables were excluded. The best model that explained the outcome was selected according to Akaike’s Information Criteria (AIC). The linearity assumption was confirmed before the inclusion of quantitative variables in bivariate and multivariate models. The seroprevalence are presented with their 95% confidence intervals adjusted by random effects (neighborhood). All statistical procedures were performed in R studio, using tidyverse, lme4 and nmet packages [29–32]

## Results

The study included 2,808 residents of five neighborhoods located in Salvador, Brazil (Fig 1); of which 1,512 were from Pau da Lima, 332 from Marechal Rondon, 367 from Alto do Cabrito, 304 from Nova Constituinte and 293 from Rio Sena (Table 1). In general, the population studied was predominantly female (58%; 1,620/2,808) and self-declared black (51%; 1,422/2,808) (Table 1).

**Table 1.**
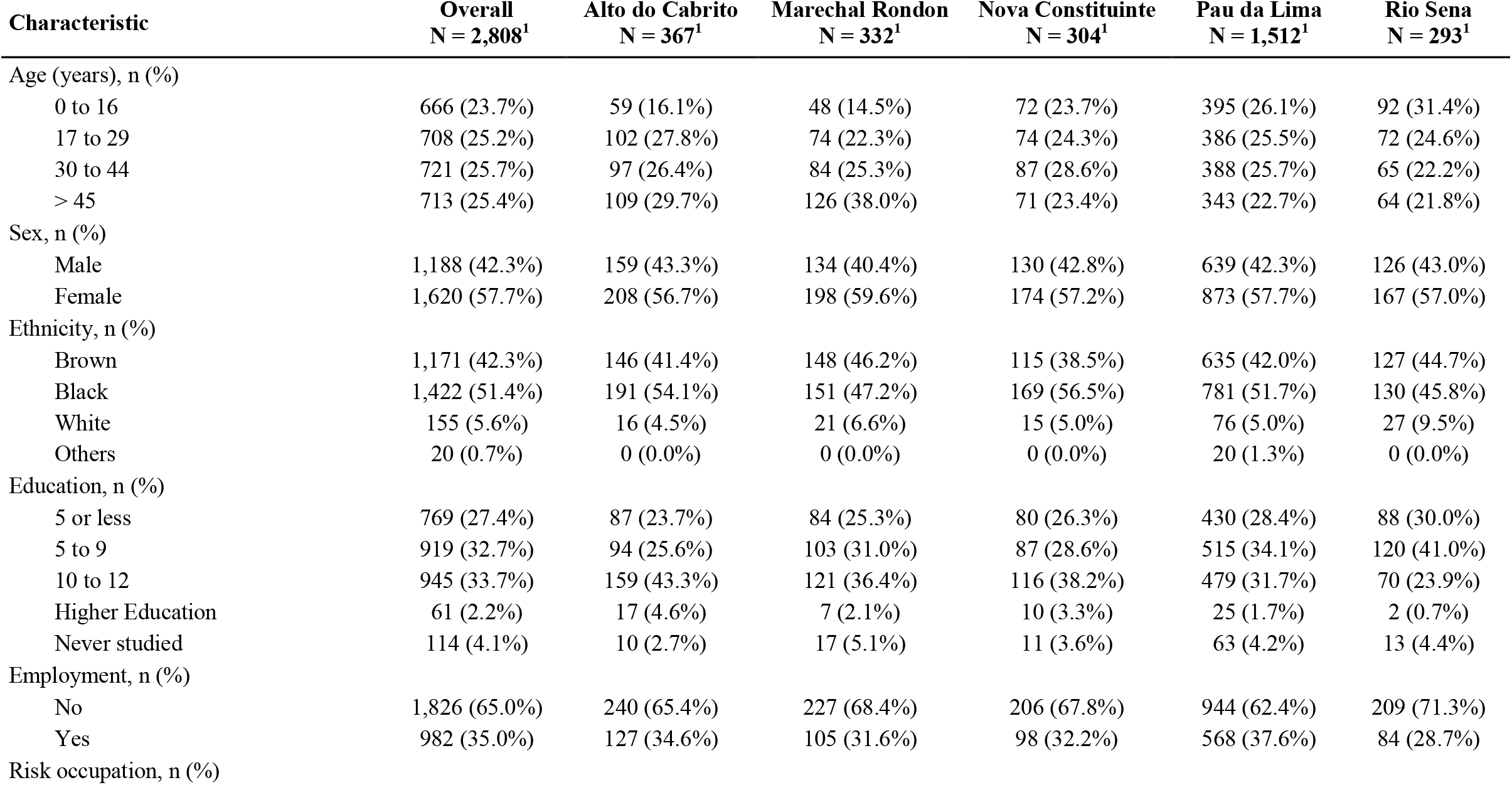
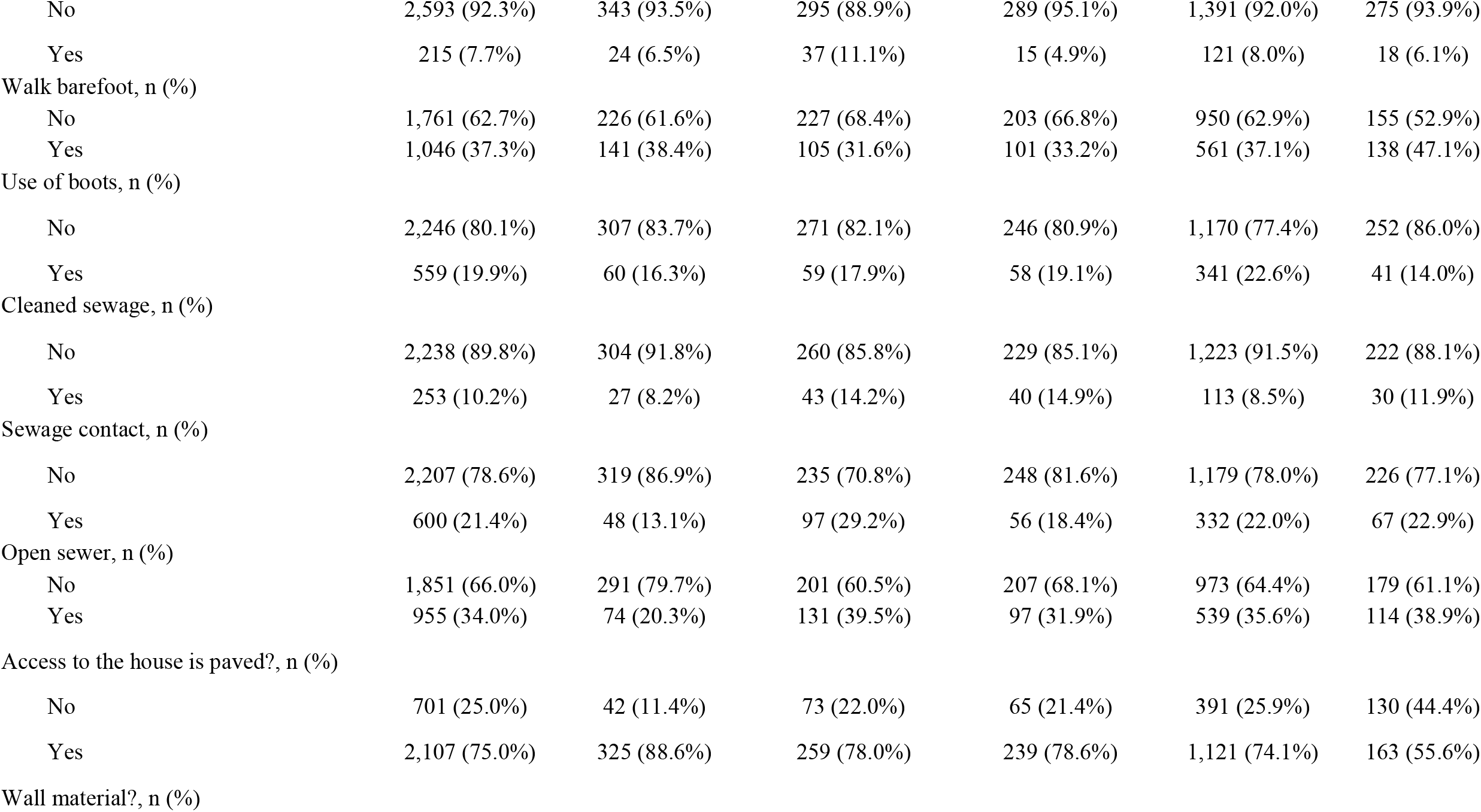
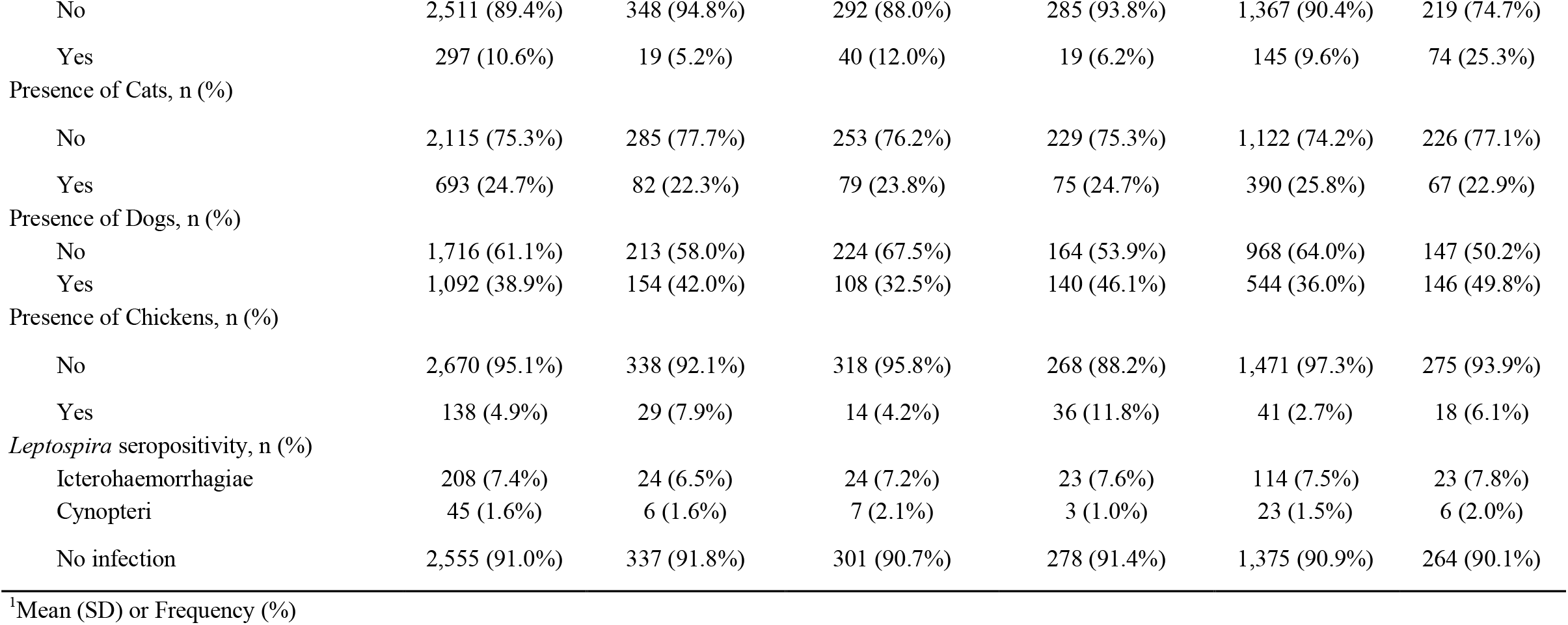
Descriptive statistics for demography and socioenvironmental variables by area.

The overall unadjusted seroprevalence was 9% (253/2,808) (95% CI 8.0% - 10.1%). Of the 253 infected, 18% (45/253) were seropositive to serogroup Cynopteri and 82% (208/253) were seropositive to serogroup Icterohaemorrhagiae. Regarding the distribution of titers, 190/208 (91%) of the samples had titers of 50 to 400, followed by 18/208 (9%) of the samples reacted to titers of 800 and 1600 for serogroup Icterohaemorrhagiae. For the Cynopteri serogroup, 43/45 (96%) of the samples had titers of 50 to 400 followed by 2/45 (4%) of the samples reacted to titers of 800 and 6400. Stratifying seroprevalence per area, the seropositivity to serogroup Icterohaemorrhagiae was lowest in AdC (6.5%) and was highest in RS (7.8%). Seroprevalence of Cynopteri ranged between 1.0% in NC and 2.1% in MR (Table 1). Seroprevalence of Icterohaemorrhagiae increased with age group (Fig 2), and females showed less chance to be infected than males (OR 0.69; 95% CI 0.52 – 0.91). This pattern was not observed for serogroup Cynopteri, which presented higher seroprevalence in the age group >45 years (Fig 2).

**Figure 2.**
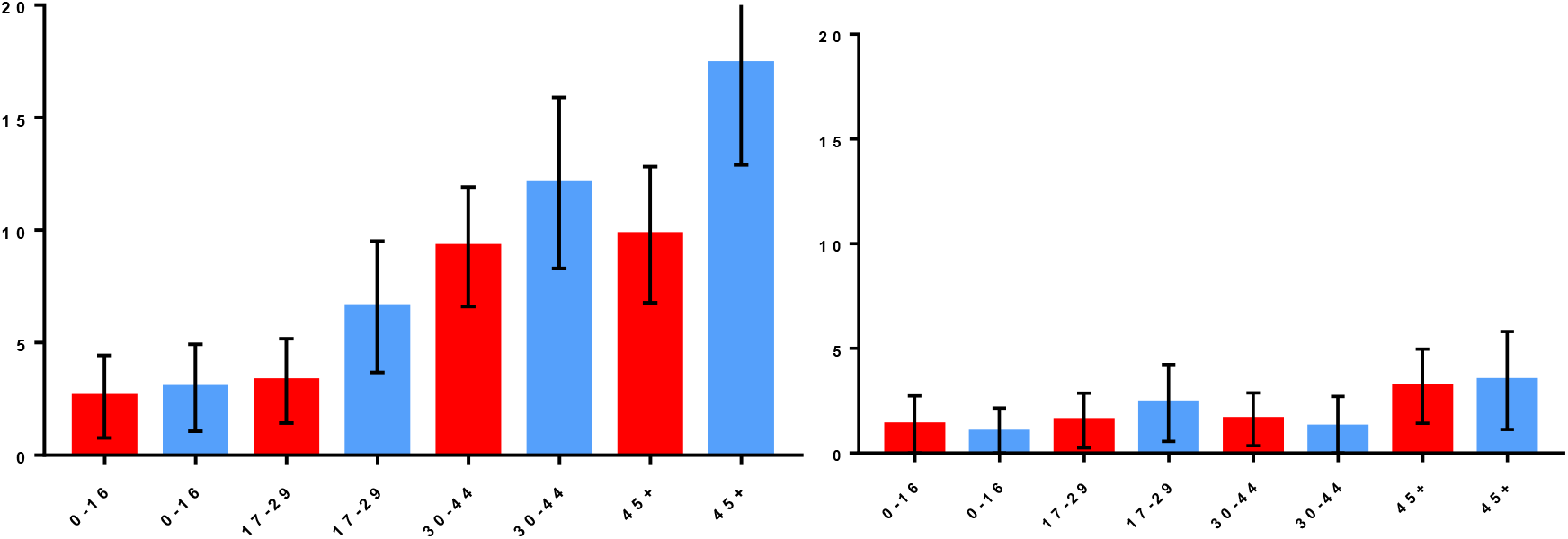
Infection rate for *Leptospira interrogans* Icterohaemorrhagiae (A) and *Leptospira kirschneri* Cynopteri (B) by gender and age. Red bars: female. Blue bars: males. Whiskers: 95% CI.

The final multivariable multinomial analysis (Table 2) which includes data for residents > 18 years of age to investigate work-related exposures (n=2,009), identified the following variables as significantly associated with the serogroup Cynopteri: age in years (as a quantitative variable) (OR 1.03; 95% CI 1.01-1.06), lives in house with unplaster walls (exposed brick) (OR 1.68; 95% CI 1.09-2.59) and presence of cats near the household (OR 2.00; 95% CI 1.03-3.88). In addition, gender (males>females (OR 1.51; 95% CI 1.09-2.10), age in years (OR 1.02; 95% CI 1.01-1.03), work-related exposures (OR 1.71; 95% CI 1.10-2.66) and contact with sewage (OR 1.42; 95% CI 1.00-2.03) were factors associated to seropositivity against Icterohaemorrhagiae.

**Table 2.**
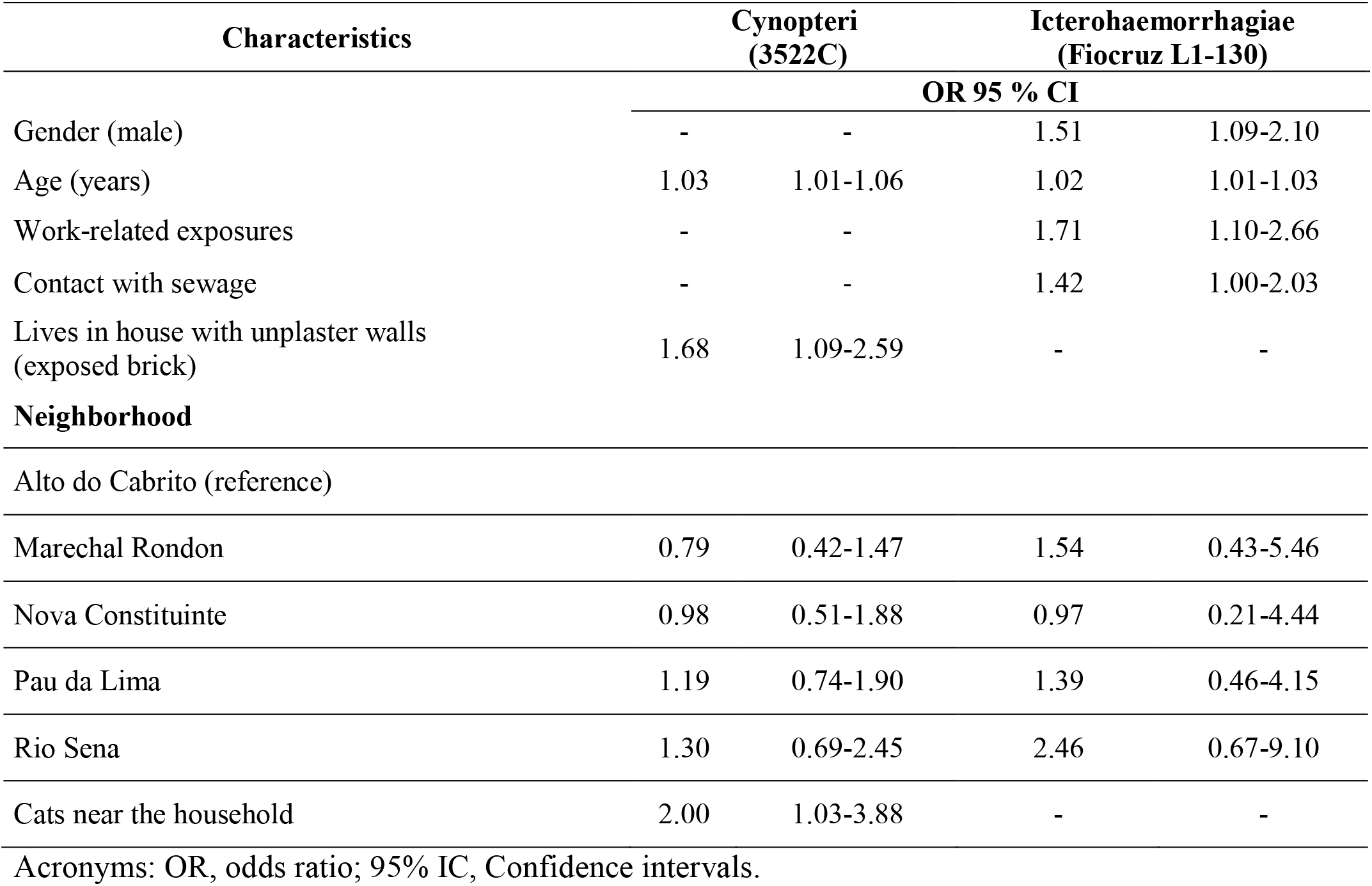
Multivariate multinomial regression of seropositivity for *Leptospira kirschneri* Cynopteri (3522C) and *Leptospira interrogans* Icterohaemorrhagiae (Fiocruz L1-130), n= 2,009). Note that there were no differences in seropositivity among neighborhoods.

Figure 3 shows differences in spatial patterns of seropositivity between serogroups Icterohaemorrhagiae and Cynopteri (yellow rectangle indicate Non-overlapping Kernel ratio concentration areas between serogroups). The objective of this complementary analysis was to verify possible overlaps in the occurrence of spatial patterns between serogroups in the territory.

**Figure 3.**
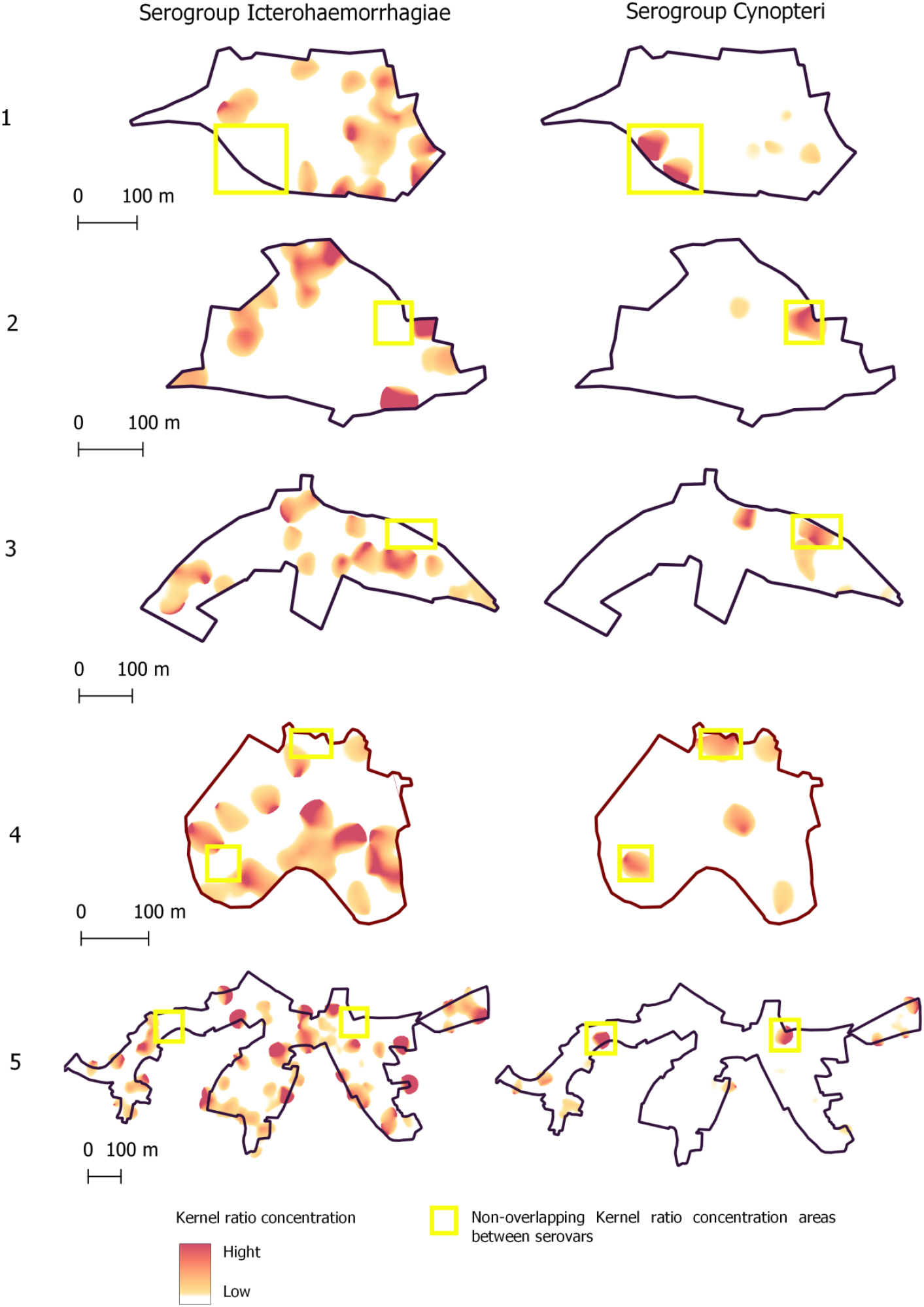
Spatial patterns of seropositivity to serogroups’ Icterohaemorrhagiae and Cynopteri (yellow rectangle) on five neighborhoods of Salvador, Bahia, Brazil. The light red-to-hard red gradient represents increasing density in smoothing analyses which used 30 meters as the bandwidth.

## Discussion

Here, we found that of *L. interrogans* serogroup Icterohaemorrhagiae was the main serogroup responsible for leptospirosis cases in Salvador [12,33]. However, we also found evidence of local exposure to *L. kirschneri* serogroup Cynopteri in residents of all communities studied. Prevalence of *L. kirschneri* serogroup Cynopteri, a serogroup considered pathogenic and for which the reservoir in an urban environment is not known, was lower than that of *L. interrogans* serogroup Icterohaemorrhagiae.

We identified differences in the demographic and socioenvironmental factors that affect seropositivity for each serogroup. Residents exposed to serogroup Cynopteri had a distinct demographic profile compared to that of serogroup Icterohaemorrhagiae. We observed higher seroprevalence for serogroup Cynopteri in ages >45 years for both genders, suggesting that behavioral differences between men and women do not affect exposure. However, for serogroup Icterohaemorrhagiae, we found that seropositivity increased with age, and males were more likely to be seropositive. This relationship between age and exposure to Icterohaemorrhagiae is similar to that observed in previous studies [16].

Occupation-related factors were associated with exposure to serogroup Icterohaemorrhagiae. Individuals who engaged in activities involving solid waste management, or contact with mud, floodwaters and/or sewage, were more likely to be seropositive. This is in line with previous results from these and similar communities, where leptospirosis is associated to work-related exposures, including occupations like subsistence farming [3]. Further, inhabitants of low-income communities are commonly enrolled in informal employment, often close to their households, highlighting the role of the environment as an important source for transmission. Interestingly, we did not detect any independent associations between work-related risk exposures and serogroup Cynopteri, maybe because have a lower number of individuals seropositive for Cynopteri.

Little is known about the animal and environmental reservoirs for serogroup Cynopteri. In our final model, we detected a positive association of individuals between anti-Cynopteri antibodies and the presence of cats in the household. Although there is a report on the occurrence of this serogroup in wild animals such as bats [34], there are also indications that cats can be exposed to *Leptospira* and higher seroprevalence of serogroup Cynopteri has been found in cats from Spain compared to other serogroups studied [35].

Among the domestic and peri-domiciliary variables, the presence of open sewage near the home was associated with greater seropositivity for serogroup Icterohaemorrhagiae. This result is consistent with a previous study [19] in which “living near an open sewer” was a factor associated with seropositivity. This further suggests that environmental exposure is an important route of indirect transmission, and it should be noted that animals other than rats can release leptospires through urine into the environment. This association has also been reported in other epidemiological studies [31,32]. In a previous study, in the same study area, it was identified that, in addition to sewage, the presence of pond water on the ground, from rainwater accumulations, can be a source of pathogenic *Leptospira* [36].

Infrastructure deficiencies in homes, for example, living in a house with unplaster walls (exposed brick), has also been considered a source of transmission for repeated exposures to *Leptospira* [19]. This may mean that risk factors for peri-domiciliary exposure may also be important for infection by serogroup Cynopteri. We believe that the structural precariousness in the household could be an indicator of low socioeconomic status, as suggested in another study [24].

It is possible that different spatial niches of reservoirs occur for serogroups Icterohaemorrhagiae and Cynopteri in urban areas, as a difference in spatial concentration patterns was observed in the analysis of the Kernel ratio (Figure 3) for the occurrence of these serogroups in the five neighborhoods evaluated. There was low overlap in human case concentration patterns for the serogroups, possibly because serogroup Icterohaemorrhagiae is associated with proximity to sewers and Cynopteri with the presence of cats, as demonstrated by statistical analysis. However, these investigations are limited by the fact that MAT is the standard test used in prevalence surveys, but this serological analysis does not actually indicate that the individual has acute disease. Additionally, we had a low prevalence of Cynopteri, which might not be possible to understand the true epidemiological pattern. Future studies may deepen to evaluate spatial correlations between the occurrence of serogroups and the abundance and mobility of rodents or felines and proximity by households.

## Conclusion

Our data show different exposure risk for different serogroups, with possible different spatial territorial niches of reservoirs, which can potentially indicate different epidemiology or transmission pathways. Further, our findings indicate cats were associated to seropositivity for serogroup Cynopteri in residents of the communities studied. Our results can serve as base for future studies in One Health approach that can elucidate the role of synanthropic, wild animals, and environment in the transmission of serogroup Cynopteri in urban areas, in addition to contributing to the orientation of public health measures to prevent future infections.

## Data Availability

The authors declare to make available all data underlying their findings without restriction in compliance with the PLOS data policy.

## Acknowledgments

The authors would like to thank the individuals from the communities who participated in this study.

## Author Contributions

Conceived and designed the experiments: FC, DO, HK, MB, AIK, MGR, YAL. Performed the experiments: FC, DO, HK, FAGP, RS, GAS, JC, FAGP, EAWJ. Analyzed the data: DO, HK, JCQ, NNJ, CGZ, ISF. Contributed reagents/materials/analysis tools: FC, DO, AIK, HK, MGR, MB, PJD, EAWJ, YAL, R, NNJ. Wrote the paper: DO, FC, MGR, HK, EAWJ, CGZ, JC, YAL, NNJ, RL, AIK, MB, FAGP, TCP.

---

**Table S1.** Bivariate models for seroprevalence of *Leptospira kirschneri* Cynopteri (3522C) and *Leptospira interrogans* Icterohaemorrhagiae (Fiocruz L1-130).

| Characteristic | Cynopteri<br>(3522C) |  |  | Icterohaemorrhagiae<br>(Fiocruz L1-130) |  |  |
| --- | --- | --- | --- | --- | --- | --- |
|  | N | OR<sup>1</sup> | 95% CI<sup>1</sup> | N | OR<sup>1</sup> | 95% CI<sup>1</sup> |
| **Individual** |  |  |  |  |  |  |
| Median Age (years) | 2,600 | 1.03 | 1.02, 1.05 | 2,763 | 1.03 | 1.02, 1.04 |
| Sex | 2,600 |  |  | 2,763 |  |  |
| Male |  | — | — |  | — | — |
| Female |  | 1.07 | 0.59, 1.99 |  | 0.69 | 0.52, 0.91 |
| Ethnicity | 2,562 |  |  | 2,724 |  |  |
| Brown |  | — | — |  | — | — |
| Black |  | 0.86 | 0.47, 1.58 |  | 0.97 | 0.72, 1.30 |
| White |  | 0.36 | 0.02, 1.75 |  | 1.21 | 0.64, 2.11 |
| Others |  | 0 |  |  | 0 |  |
| Education | 2,600 |  |  | 2,763 |  |  |
| 5 or less |  | — | — |  | — | — |
| 5 to 9 |  | 0.6 | 0.26, 1.34 |  | 1.09 | 0.76, 1.57 |
| 10 to 12 |  | 0.75 | 0.35, 1.61 |  | 0.95 | 0.66, 1.38 |
| Higher Education |  | 0.86 | 0.05, 4.40 |  | 0.43 | 0.07, 1.43 |
| Never studied |  | 3.64 | 1.35, 8.98 |  | 1.43 | 0.69, 2.73 |
| Employment | 2,600 |  |  | 2,763 |  |  |
| No |  | — | — |  | — | — |
| Yes |  | 0.87 | 0.45, 1.62 |  | 1.73 | 1.30, 2.29 |
| Risk occupation | 2,600 |  |  | 2,763 |  |  |
| No |  | — | — |  | — | — |
| Yes |  | 0.96 | 0.23, 2.67 |  | 2.72 | 1.81, 3.99 |
| **Risk exposures** |  |  |  |  |  |  |
| Walk barefoot | 2,599 |  |  | 2,762 |  |  |
| No |  | — | — |  | — | — |
| Yes |  | 0.75 | 0.39, 1.39 |  | 0.88 | 0.65, 1.18 |
| Use of boots | 2,597 |  |  | 2,760 |  |  |
| No |  | — | — |  | — | — |
| Yes |  | 0.75 | 0.30, 1.58 |  | 1.22 | 0.86, 1.69 |
| Cleaned sewage | 2,286 |  |  | 2,448 |  |  |
| No |  | — | — |  | — | — |
| Yes |  | 1.3 | 0.44, 3.05 |  | 2.55 | 1.75, 3.65 |
| Sewage contact | 2,599 |  |  | 2,762 |  |  |
| No |  | — | — |  | — | — |
| Yes |  | 1.38 | 0.68, 2.62 |  | 1.4 | 1.01, 1.92 |
| Open sewer | 2,598 |  |  | 2,761 |  |  |
| No |  | — | — |  | — | — |
| Yes |  | 1.32 | 0.71, 2.40 |  | 1.26 | 0.94, 1.69 |
| Access to the house is paved? | 2,600 |  |  | 2,763 |  |  |
| No |  | — | — |  | — | — |
| Yes |  | 1.03 | 0.54, 2.14 |  | 1.03 | 0.74, 1.44 |
| Wall material? | 2,600 |  |  | 2,763 |  |  |
| No |  | — | — |  | — | — |
| Yes |  | 1.1 | 0.38, 2.57 |  | 1.6 | 1.06, 2.36 |
| **Presence of animals in the household** |  |  |  |  |  |  |
| Presence of Cats | 2,600 |  |  | 2,763 |  |  |
| No |  | — | — |  | — | — |
| Yes |  | 1.89 | 1.01, 3.44 |  | 1.09 | 0.78, 1.50 |
| Presence of Dogs | 2,600 |  |  | 2,763 |  |  |
| No |  | — | — |  | — | — |
| Yes |  | 0.86 | 0.45, 1.57 |  | 0.94 | 0.70, 1.25 |
| Presence of Chickens | 2,600 |  |  | 2,763 |  |  |
| No |  | — | — |  | — | — |
| Yes |  | 1.46 | 0.35, 4.09 |  | 1.71 | 0.96, 2.85 |

<sup>1</sup>OR = Odds Ratio, CI = Confidence Interval

## References

1. Ko AI, Goarant C, Picardeau M. Leptospira: the dawn of the molecular genetics era for an emerging zoonotic pathogen. Nat Rev Microbiol. 2009;7: 736–747. doi:10.1038/nrmicro2208

2. Bharti AR, Nally JE, Ricaldi JN, Matthias MA, Diaz MM, Lovett MA, et al. Leptospirosis: A zoonotic disease of global importance. Lancet Infect Dis. 2003;3: 757–771. doi:10.1016/S1473-3099(03)00830-2

3. Mcbride AJA, Athanazio DA, Reis MG, Ko AI. Leptospirosis. Trop Travel Dis. 2005;18: 376–386.

4. Costa F, Hagan JE, Calcagno J, Kane M, Torgerson P, Martinez-Silveira MS, et al. Global morbidity and mortality of leptospirosis: a systematic review. PLoS Negl Trop Dis. 2015; 1–19. doi:10.1371/journal.pntd.0003898

5. Goarant C. Leptospirosis: risk factors and management challenges in developing countries. Res Rep Trop Med. 2016;Volume 7: 49–62. doi:10.2147/rrtm.s102543

6. Bierque E, Thibeaux R, Girault D, Soupé-Gilbert ME, Goarant C. A systematic review of Leptospira in water and soil environments. PLoS One. 2020;15:1–22. doi:10.1371/journal.pone.0227055

7. Picardeau M. Virulence of the zoonotic agent of leptospirosis: Still terra incognita? Nat Rev Microbiol. 2017;15: 297–307. doi:10.1038/nrmicro.2017.5

8. Vincent AT, Schiettekatte O, Goarant C, Neela VK, Bernet E, Thibeaux R, et al. Revisiting the taxonomy and evolution of pathogenicity of the genus Leptospira through the prism of genomics. PLoS Negl Trop Dis. 2019;13: e0007270. doi:10.1371/journal.pntd.0007270

9. Casanovas-Massana A, Hamond C, Santos LA, de Oliveira D, Hacker KP, Balassiano I, et al. Leptospira yasudae sp. Nov. and Leptospira stimsonii sp. nov., two new species of the pathogenic group isolated from environmental sources. Int J Syst Evol Microbiol. 2019;70: 1450–1456. doi:10.1099/ijsem.0.003480

10. Picardeau M. Diagnosis and epidemiology of leptospirosis. Médecine Mal Infect. 2013;43: 1–9. doi:10.1016/S0992-5945(17)30102-2

11. Faine S, Adler B, Bolin C, Perolat P. Leptospira and Leptospirosis. 2^a^ ed. MediSci. Melbourne, Australia; 1999.

12. Ko AI, Reis MG, Dourado CMR, Johnson Jr WD, Riley LW. Urban epidemic of severe leptospirosis in Brazil. Lancet. 1999;354: 820–825. Available: http://www.ncbi.nlm.nih.gov/pubmed/10485724

13. Hacker KP, Sacramento GA, Cruz JS, De Oliveira D, Nery N, Lindow JC, et al. Influence of rainfall on leptospira infection and disease in a tropical urban setting, Brazil. Emerg Infect Dis. 2020;26: 311–314. doi:10.3201/eid2602.190102

14. Trevejo RT, Rigau-Pérez JG, Ashford DA, McClure EM, Jarquín-González C, Amador JJ, et al. Epidemic Leptospirosis Associated with Pulmonary Hemorrhage—Nicaragua, 1995. J Infect Dis. 1998;178: 1457–1463. doi:10.1086/314424

15. Amilasan AST, Ujiie M, Suzuki M, Salva E, Belo MCP, Koizumi N, et al. Outbreak of leptospirosis after flood, the Philippines, 2009. Emerg Infect Dis. 2012;18: 91–94. doi:10.3201/eid1801.101892

16. Reis RB, Ribeiro GS, Felzemburgh RDM, Santana FS, Mohr S, Melendez Axto, et al. Impact of environment and social gradient on Leptospira infection in urban slums. PLoS Negl Trop Dis. 2008;2: 11–18. doi:10.1371/journal.pntd.0000228

17. UN-HABITAT. Streets as public spaces and drivers of urban prosperity. 2013.

18. UN-HABITAT. State of the World’s Cities 2010/2011: Bridging The Urban Divide. In: Earthscan, GB. 2010 p. 224.

19. Santos M. A urbanização brasileira. Ed. Hucite. São Paulo; 1993.

20. Spink MJP. Viver em áreas de risco: reflexões sobre vulnerabilidades socioambientais. EDUC: Terc. São Paulo; 2018.

21. Felzemburgh RDM, Ribeiro GS, Costa F, Reis RB, Hagan JE, Melendez Axto, et al. Prospective Study of Leptospirosis Transmission in an Urban Slum Community: Role of Poor Environment in Repeated Exposures to the Leptospira Agent. PLoS Negl Trop Dis. 2014;8: 1–9. doi:10.1371/journal.pntd.0002927

22. Hagan JE, Moraga P, Costa F, Capian N, Ribeiro GS, Wunder EA, et al. Spatiotemporal determinants of urban leptospirosis transmission: Four-year prospective cohort study of slum residents in Brazil. PLoS Negl Trop Dis. 2016;10: 1–16. doi:doi:10.1371/journal.pntd.0004275

23. Costa F, Porter FH, Rodrigues G, Farias H, de Faria MT, Wunder E a, et al. Infections by Leptospira interrogans, Seoul virus, and Bartonella spp. among Norway rats (Rattus norvegicus) from the urban slum environment in Brazil. Vector borne zoonotic Dis. 2014;14: 33–40. doi:10.1089/vbz.2013.1378

24. Costa F, Ribeiro GS, Felzemburgh RDM, Santos N, Reis RB, Santos AC, et al. Influence of Household Rat Infestation on Leptospira Transmission in the Urban Slum Environment. PLoS Negl Trop Dis. 2014;8: 1–8. doi:10.1371/journal.pntd.0003338

25. Khalil H, Santana R, de Oliveira D, Palma F, Lustosa R, Eyre MT, et al. Poverty, sanitation, and leptospira transmission pathways in residents from four Brazilian slums. PLoS Negl Trop Dis. 2021;15: 1–15. doi:10.1371/journal.pntd.0009256

26. Silverman B. Estimativa de densidade para estatísticas e análise de dados. Chapman & Hall, Londres; 1986.

27. Gómez Villafañe IE, Muschetto E, Busch M. Movements of Norway rats (Rattus norvegicus) in two poultry farms, Exaltación de la Cruz, Buenos Aires, Argentina. Mastozoología Neotrop. 2008;15: 203–208.

28. Hagan JE, Moraga P, Costa F, Capian N, Ribeiro GS, Wunder EA, et al. Spatiotemporal Determinants of Urban Leptospirosis Transmission: Four-Year Prospective Cohort Study of Slum Residents in Brazil. PLoS Negl Trop Dis. 2016;10: 1–16. doi:10.1371/journal.pntd.0004275

29. RStudio: Integrated Development for R. RStudio, PBC, Boston M. RStudio Team. 2020. Available: http://www.rstudio.com/

30. Team RC. R: A language and environment for statistical computing. Foundation for Statistical Computing. Vienna, Austria; 2019. Available: https://www.r-project.org/

31. Venables W, Ripley B. Modern Applied Statistics with S. New York; 2002. Available: https://www.stats.ox.ac.uk/pub/MASS4/

32. D B, M M, B B, S W. Fitting Linear Mixed-Effects Models Using lme4. J Stat Softw. 2015;67: 1–48. doi:10.18637/jss.v067.i01

33. Barocchi MA, Ko AI, Ferrer SR, Faria MT, Reis MG, Riley LW. Identication of New Repetitive Element in Leptospira interrogans Serovar Copenhageni and Its Application to PCR-Based Differentiation of Leptospira Serogroups. Clin Microbiol. 2001;39: 191–195. doi:10.1128/JCM.39.1.191

34. Matthias MA, Campos KJ, Calderon M, Willig MR, Pacheco V, Gotuzzo E, et al. Diversity of bat-associated Leptospira in the peruvian amazon inferred by bayesian phylogenetic analysis of 16s ribosomal DNA sequences. Am Soc Trop Med Hyg Divers. 2005;73: 964–974.

35. Murillo A, Cuenca R, Serrano E, Marga G, Ahmed A, Cervantes S, et al. Leptospira detection in cats in Spain by serology and molecular techniques. Int J Environ Res Public Health. 2020;17. doi:10.3390/ijerph17051600

36. Casanovas-Massana A, Costa F, Riediger IN, Cunha M, Oliveira D de, Mota DC, et al. Spatial and temporal dynamics of pathogenic Leptospira in surface waters from the urban slum environment. Water Res. 2018;130: 176–184. doi:10.1016/j.watres.2017.11.068

